# Pandemic Politics: Timing State-Level Social Distancing Responses to COVID-19

**DOI:** 10.1101/2020.03.30.20046326

**Authors:** Christopher Adolph, Kenya Amano, Bree Bang-Jensen, Nancy Fullman, John Wilkerson

## Abstract

Social distancing policies are critical but economically painful measures to flatten the curve against emergent infectious diseases. As the novel coronavirus that causes COVID-19 spread throughout the United States in early 2020, the federal government issued social distancing recommendations but left to the states the most difficult and consequential decisions restricting behavior, such as canceling events, closing schools and businesses, and issuing stay-at-home orders. We present an original dataset of state-level social distancing policy responses to the epidemic and explore how political partisanship, COVID-19 caseload, and policy diffusion explain the timing of governors’ decisions to mandate social distancing. An event history analysis of five social distancing policies across all fifty states reveals the most important predictors are political: all else equal, Republican governors and governors from states with more Trump supporters were slower to adopt social distancing policies. These delays are likely to produce significant, on-going harm to public health.

## Introduction

Social distancing, or practices which reduce the probability of contact between infected and non-infected people, has emerged as the primary tool for reducing the spread of the novel coronavirus that causes COVID-19.(1; 2; 3) As of March 2020, other methods for fighting infectious disease are mostly unavailable or ineffective due to the characteristics of this pathogen

1. it is an emergent virus to which there is no pre-existing immunity, available vaccine, or proven treatment;
2. it spreads easily through human contact and airborne droplets, leading to exponential growth in cases; and
3. those affected may be contagious during a prolonged asymptomatic incubation period, and many never develop symptoms distinctive from a mild flu, making it difficult to identify and isolate the infected before they pass the virus to others.(4)

Broad public mandates for social distancing are also vital to help manage a key multiplier of COVID-19 fatality rates: a substantial fraction of patients require lengthy (on average 8 bed days) intensive care unit (ICU) support to survive acute respiratory distress, and uncontrolled spread of the disease is expected to generate at its peak far greater demand for ICU care than existing capacity can meet.(5; 2; 6) Public health experts have pleaded with officials to quickly mandate social distancing to flatten the curve of coronavirus infections – that is, reduce the peak caseload – before exponential case growth leads to hundreds of thousands or millions of deaths. In particular, it is important to keep peak levels low to prevent the fatality rate from rising to the levels seen in areas where hospitals are overwhelmed with cases.(7)

In the United States, the emergence of the coronavirus has created a natural experiment in which elected officials face incredibly urgent and far-reaching policy decisions for which they typically have no personal experience or expertise, and for which the history of policy examples is either mere days old, or so dated as to be largely unknown except by public health experts. The most significant policy decisions of this kind – both in terms of their potential to mitigate the epidemic and their economic consequences – are mandates for social distancing through restrictions on public gatherings; closure or restriction of schools, restaurants, and other non-essential businesses; and orders for the public to stay at home.(8) Under the federal and state constitutions, the key actions fall to executives, and particularly to governors, whose powers in public health emergencies are typically singular and extensive.

In particular, the exponential growth of cases of an emergent virus has unique implications for policy making. Estimates of the uncontrolled doubling-time of COVID-19 cases vary and are complicated by the slow rollout of effective testing in the US, but many studies find doubling times in the range from four to seven days.(9; 10; 3; 2) These estimates correspond to alarming growth rates of 10.4% to 26.0% per day. Even if the true doubling time of infections prior to social distancing is as long as a week, delaying these measures just three days would, all else equal, raise the eventual peak number of cases by more than 30%, potentially increasing total deaths by thousands or more in a given state.

Evidence from the 1918 influenza pandemic underscores the stakes for elected officials. In that earlier natural experiment, American cities faced similar decisions regarding social distancing policies. For example, Philadelphia decided to hold a parade welcoming soldiers returning from World War I, while St. Louis cancelled its parade, and ultimately experienced one eighth as many deaths per capita during that wave of influenza. The height of the infection in Saint Louis was two months later than in Philadelphia, and was far lower, with deaths peaking at fifty per hundred thousand rather than 250 per hundred thousand.(11; 12) Shortly after community transmission of the coronavirus was discovered in Washington and California in late February 2020, public health experts drew the comparison between Philadelphia and Saint Louis, hoping to inspire policymakers to promote social distancing. Nevertheless, in an echo of the decisions made a century before, there are significant differences in the timing of governors’ decisions to mandate social distancing. For instance, on 6 March 2020, after Kentucky had its first confirmed case of COVID-19, Governor Andy Beshear (D) immediately called a state of emergency, encouraged social distancing, and closed bars and restaurants ten days later. But when neighboring Tennesse uncovered its first confirmed case of COVID-19 on March 5th, Governor Bill Lee (R) waited until 12 March to declare a state of emergency and finally closed restaurants and bars on 22 March.(13)

Why did governors’ responses to a sudden, shared threat differ? Under normal circumstances, governors’ policy decisions are shaped by the national leaders and agenda of their political party and the demands of their own constituents, particularly those in their partisan base. Governors may also look to innovative peers and neighbors for examples to follow. But the fast-moving nature of the coronavirus threat, the need to rely on experts to understand complex scientific material, and uncertainty about the nature and scope of the threat might scramble all or some of these usual motivations.

The public health case for mandatory social distancing in the United States developed quickly in the wake of the first reports of community transmission on 26 February in Washington state. One might expect social distancing measures to quickly follow across the fifty states, perhaps more quickly in states with greater number of confirmed cases. But mandating social distancing is a difficult decision for any political leader. If these measures are successful in preventing widespread mortality, many members of the public who suffered the costs of these mandates may never fully comprehend the benefits. Indeed, the more successful the intervention, the more likely it will appear to many as an “overreaction.” Moreover, some states faced higher potential costs from these policies: closing schools is more painful in states where more children depend on schools for subsidized lunches and shutting restaurants and public places is more difficult in states highly dependent on tourism.(14) More generally, states with more limited economic resources are likely far less able to weather the deprivations caused by economic shutdowns, or to expand social insurance protections during mandated social distancing, and thus may be considerably more reluctant to take these steps.

But the greatest barrier to swift social distancing measures seems political. Numerous surveys have found significant partisan divides in public opinion about the severity of the coronavirus threat.(15; 16) Countering the message from public health leaders, the White House downplayed COVID-19. On 4 March 2020, President Trump insisted that COVID-19 was similar to the flu; two days later, he falsely claimed the situation in Italy was improving, and that the US was handling coronavirus much better than other industrialized countries. As late as 15 March 2020, with reported cases rising rapidly, Trump still maintained that the epidemic within the US was under control.(17) Similar messages filled news sources relied on by Republican voters. Fox Business host Trish Reagan insisted the pandemic warnings were a Democratic hoax and another effort to impeach the President, while Sean Hannity, another Fox News personality, validated the conspiracy theory that coronavirus was an effort by the “deep state” to “manipulate markets, suppress dissent and push mandated medicines.”(18) For governors already reluctant to impose draconian measures, this messaging both provided cover for inaction and may have reduced public support, particularly among Republican voters. And as social distancing measures have spread, Republican leaders, including Trump, have been more vocal in their desire to quickly rollback these measures to restore the economy.

Moving beyond general impressions of the determinants of social distancing poses challenges: States with Democratic governors and voters tend to be both richer than Republican states, and to be more urban and coastal. The seemingly quicker action of “blue states” could simply be a result of the coronavirus first emerging in cities like Seattle, San Francisco, and New York, as well as in states better able to endure prolonged social distancing mandates. Likewise, disentangling the role of Republican governors from Republican voters is challenging, though some blue states, like Maryland, have Republican governors (and vice versa).

To investigate why some states acted quickly to implement social distancing while others took longer, we identified five critical areas of social distancing policy – recommendations and restrictions against public gatherings, mandatory school closures, restrictions on the normal operation of restaurants, closure of non-essential businesses, and stay-at-home orders – and collected the dates each state first announced such measures. Our primary objective is to explain the variation in timing of these measures across states using an event history analysis. This approach allows us to statistically separate the impact of having a Republican governor or a Republican electorate from a series of potential confounders, including the number of confirmed COVID-19 cases in the state, state income, and cases and policies in neighboring states.

Our findings are unambiguous: political variables are the strongest predictor of the early adoption of social distancing policies. All else equal, states with Republican governors and Republican electorates delayed each social distancing measure by an average of 2.70 days (95% CI: 2.49 to 2.88), a far larger effect than any other factor, including state income per capita, the percentage of neighboring states with mandates, or even confirmed cases in each state.

### State-Level Social Distancing Measures

We study state social distancing measures enacted over the period from the first reported case of transmission in the US on 26 February 2020 up through 23 March 2020, at which point all states had at least one social distancing policy.^1^ To capture variation in these policies across states over time, we draw on data compiled by the National Governors’ Association (NGA), which we verified and further documented by collecting information directly from state government websites.(20) We focus here on five measures

1. Recommendations or restrictions on gatherings: We code the date on which the first such measure was announced, regardless of the size of gathering specified.^2^
2. School closures: We code the date that the governor announces formal closure of public K-12 schools.
3. Restaurant restrictions: We code the date on which states first announced mandatory restrictions on in-person dining, including maximum capacity limits likely to render most restaurants non-viable.
4. Non-essential business closures: This coding does not distinguish among differences in state classifications of essential and non-essential businesses, which differ by state.
5. Stay-at-home orders: This coding includes mandates to stay at home but not advisory orders that recommend that citizens remain at home.

Although we also coded implementation dates for each of these policies, our focus is on the dates policies were announced, for two reasons. First, we expect much of the effect of these emergency measures on social behavior to be immediate. The state’s power to compel socially responsible behavior is often said to act through “quasi-voluntary compliance,” in which most citizens choose pro-social behaviors given a cue from the state that is backed with the threat of sanction.(21) This effect should be all the stronger to the extent people treat costly social distancing measures as a credible signal of the viral threat. Second, our focus is on predicting the timing of governor’s actions, which naturally points to announcement dates, rather than implementation dates which may be delayed (in the case of school closures, for example), by weekends and spring breaks.

Figure 1 displays the timing of different states’ adoptions of these five social distancing policies over the four-week study period. No state acted before 10 March, a full thirteen days after the first report of community transmission, yet by 15 March, uptake was rapid across states. In general, states typically started social distancing through recommendations or restrictions on public gatherings, then turned to restrictions on schools and restaurants. Finally, a minority of states have added closures of non-essential business and stay-at-home orders. By the end of our study period, all states except Arkansas and Mississippi had recommended against or restricted public gatherings, 44 states had closed schools and restricted restaurant operations, nineteen states closed non-essential businesses, and fourteen states issued stay-at-home orders. The final plot on the bottom right highlights the rapid growth in states enacting multiple social distancing measures, as well as the presence of a substantial number of laggard states still lacking most social distancing policies as late as 23 March 2020.

**Figure 1.**
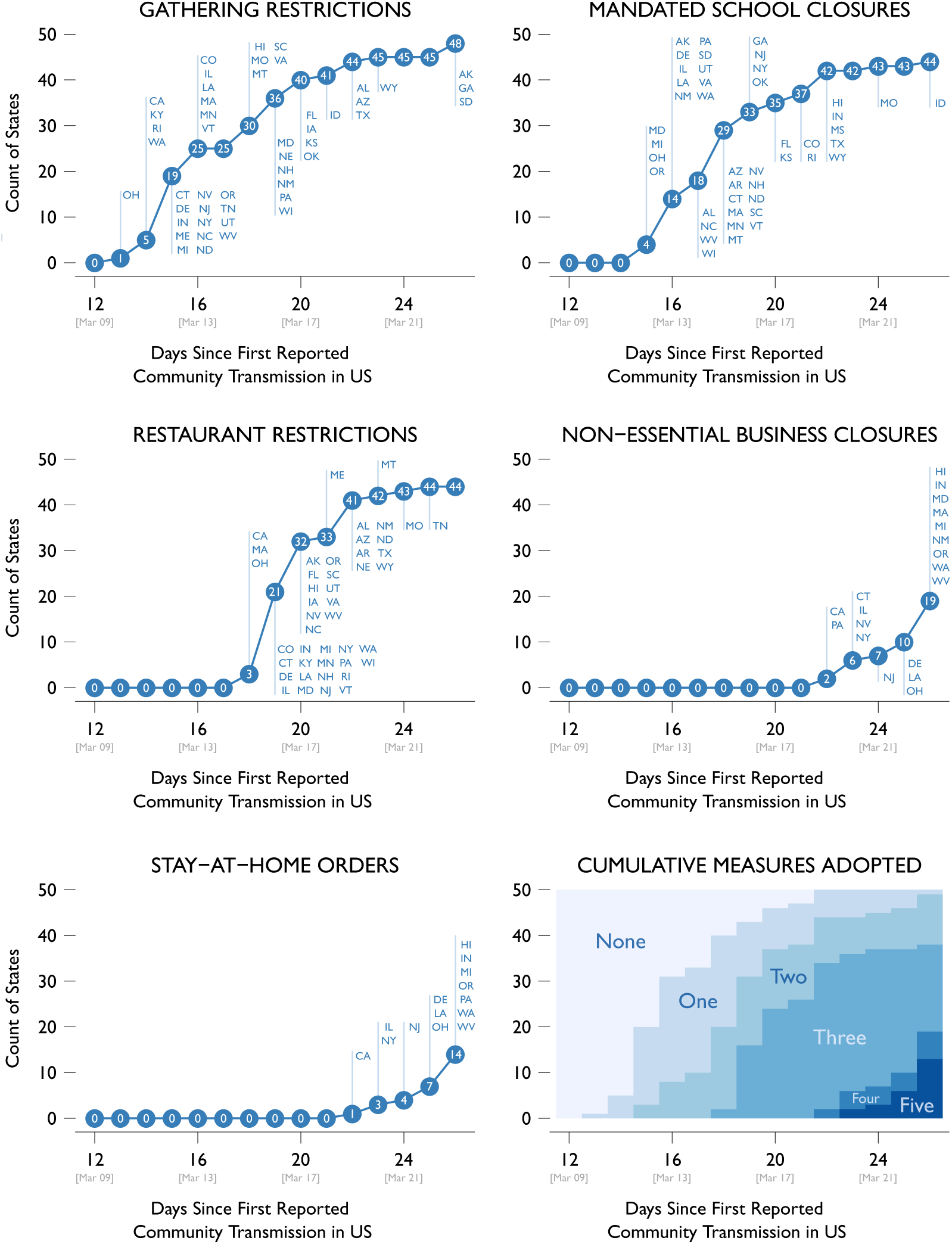
The diffusion of five social distancing measures across the states through 23 March 2020. States recorded by date of policy announcement. Sources: National Governors Association and authors’ original data collection. Data available at http://covid19statepolicy.org/

### Modeling Social Distancing Policy with Event History Analysis

We estimate an event history model to predict the timing of implementing social distancing directives across U.S. states from 26 February 2020 to 23 March 2020.^3^ We model the likelihood that a state will implement each social distancing policy as a function of time (measured in days) with a pooled, stratified Cox proportional hazards model, often referred to as the Wei-Lin-Weissfeld marginal model.(22) In our application, this approach allows us to examine the common factors affecting implementation of social distancing across states by 1) pooling the five social distancing measures shown in Figure 1 in a single model, (2) stratifying baseline hazards across the five policy types to allow for varying underlying tendencies to adopt some policies more quickly than others, and (3) clustering standard errors by state.

As usual, the Cox model allows us to estimate a baseline hazard rate, which shifts in proportion to changes in covariates, and accommodates right-censored cases (states that have not yet adopted a given social distancing measure at the end of the study period). The baseline hazard rate also captures any purely national trends, such as the common tendency of states to adopt social distancing policies as national deaths climb or public awareness of COVID-19 increases, while leaving cross-state variation to be explained by covariates.

We expect state level responses to vary based on differences in social, economic and political costs. We include five covariates in our baseline model

#### The number of confirmed cases of COVID-19 in the state

While confirmed cases of COVID-19 are undercounts of actual cases, confirmed cases were the only data available to officials making decisions in real time.(23) We explored a variety of alternative

#### Gross state product (GSP) per capita

This variable captures the ability of richer states to better withstand the impact of business shutdowns, support small businesses and schools through closure, and maintain social safety nets.(24) The variable enters the model logged to account for diminishing returns to greater wealth.

#### Republican Governor

A binary variable indicating the presence of a Republican, as opposed to a Democratic, governor.(25) Governors have extensive unchecked emergency powers, which they may exercise in accordance with their political incentives and ideology regardless of the broader partisan composition of the state.

#### Percent of voters choosing Trump in 2016

A proxy for general support for the Republican party within the state.(26) Although governors have the power to act independently in a public health crisis, they may take into account the electoral consequences of those actions, especially if they are likely to be unpopular with the dominant partisan base in their state.

#### Percentage of neighboring states enacting each social distancing measure

This variable captures the degree of policy diffusion, or states learning from their neighbors’ responses to a common policy problem.

In addition to these covariates, we explored a variety of alternative explanatory factors. These included the count of COVID-19 deaths within the state,(23) the total number of confirmed COVID-19 cases in neighboring states, Fox News viewers as a percentage of the population,(27) the spread of social distancing policies among the states from which each state typically borrows policies,(28) the percentage of state employment dependent on tourism,(29) the percentage of state residents who were at least 70 years of age,(30) the percentage of school children receiving reduced price lunches,(31) and the percentage of state residents with a college degree.(32) None of these variables had substantively and statistically significant relationships with social distancing measures when added to our baseline model, nor did they substantially alter the results of the baseline model.

## Results

We review the determinants of delays in state-level COVID-19 social distancing measures in order of substantive impact, quantifying the impact of each factor in two ways. First, we show the degree to which each factor – such as governors’ partisanship, the presence of Trump voters, or actions by neighboring states – reduces the chance a state acts to impose a new social distancing mandate on a given date (Figure 2; see Appendix for tabular results). This is the well-known hazard ratio, a common summary of event history models. Second, we use our estimated model to simulate the average delay each factor would cause if it were present in each state. That is, we might ask: if every state had a Republican governor, how many days later would they have implemented each social distancing measure than if every state had a Democratic governor. This is known as an average marginal effect and helps quantify our model results in policy-relevant terms (Figure 3).^4^

**Figure 2.**
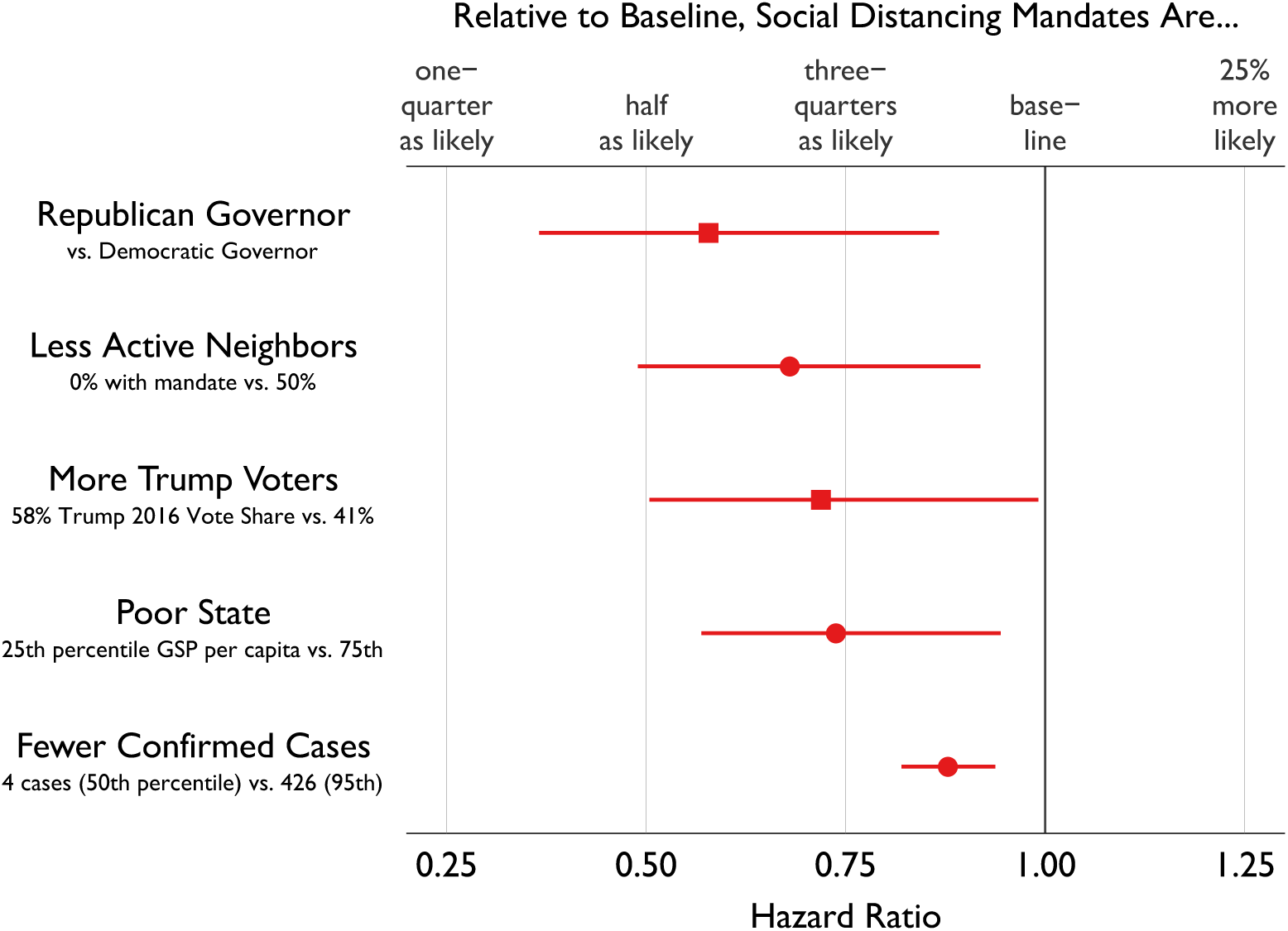
Relative probability of adopting an additional social distancing measure, by factor. Estimated hazard ratios obtained from a pooled stratified Cox proportional hazards model on all social distancing policies enacted by the fifty states, 26 February – 23 March 2020. Squares mark results related to partisanship. See Table 1 for additonal model details.

**Figure 3.**
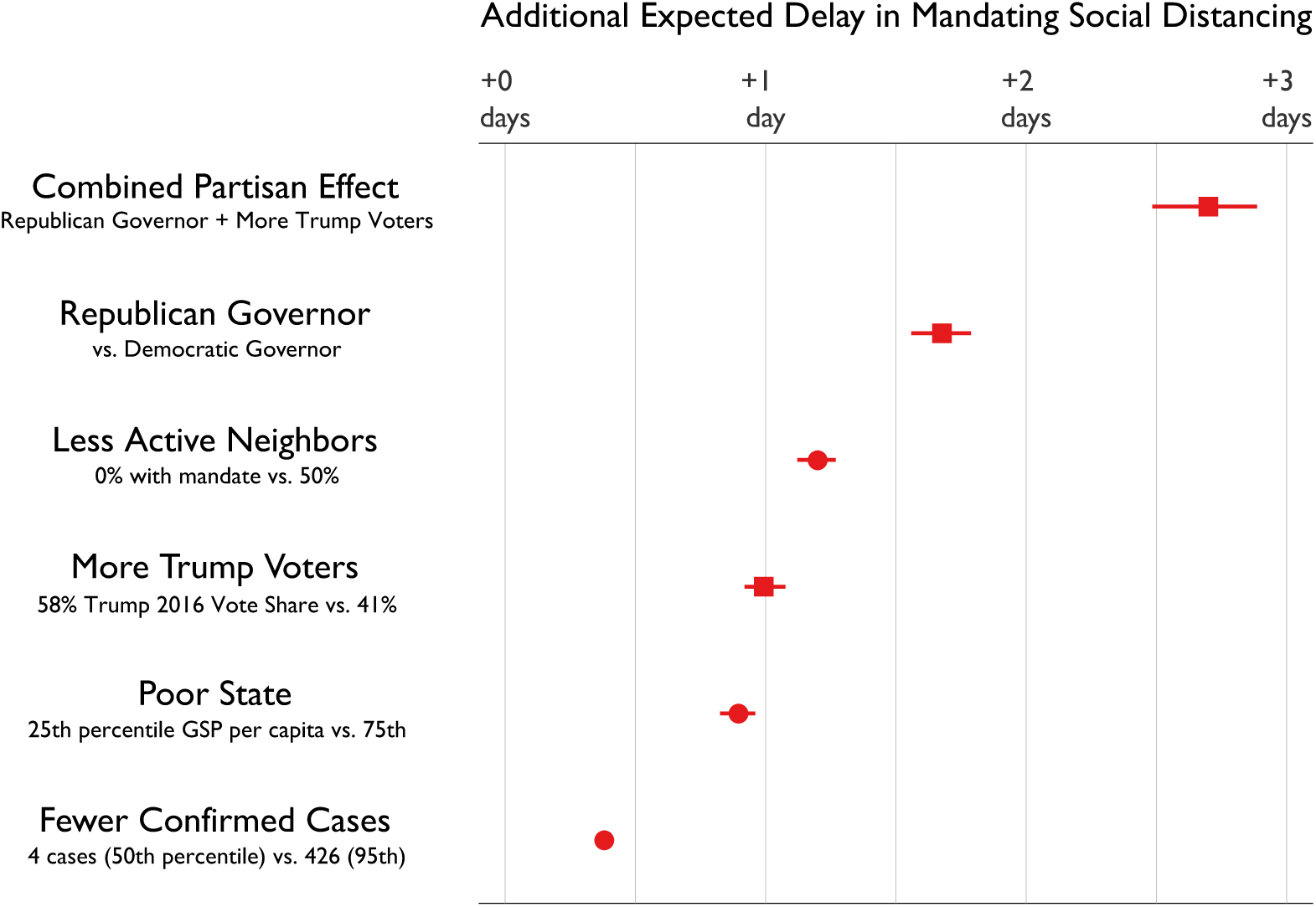
Expected delay in adopting an additonal social distancing measure, by factor. Estimated average marginal effects obtained by post-estimation simulation from a pooled stratified Cox proportional hazards model on all social distancing policies enacted by the fifty states, 26 February – 23 March 2020. Confidence intervals are bootstrapped using non-parametric step functions. Squares mark results related to partisanship. See Table 1 for additonal model details.

### Republican-leaning states are slower to adopt social distancing policies

At any given time within the study period, Republican governors were 42.2% (95% CI: 13.5 to 63.2) less likely to mandate social distancing than their Democratic counterparts. Holding constant other variables – including the 2016 Trump vote share – at their observed values in each state, on average, Republican governors delay each state-level social distancing measure by 1.68 days (95% CI: 1.57 to 1.78). At the same time, holding constant the Governor’s party affiliation, states with more Trump voters were less likely to adopt social distancing. A state at the 75th percentile of 2016 Trump vote share was 28.1% (95% CI: 1.1 to 49.3) less likely to adopt an additional mandate at a given time when compared to a state at the 25th percentile of Trump support, which resulted in an average delay of 0.99 days (95% CI: 0.93 to 1.07). Republican governors often go together with Trump-voting electorates; on average, these states endured a combined partisan delay of 2.70 days (95% CI: 2.49 to 2.88). Barring positive developments in the fight against COVID-19, the public health impact of this delay is likely to be massive: in a state where coronavirus infections are doubling every seven days, this would raise the peak caseload by 30.6%. In a state where infections are doubling every three days, Republican partisanship might raise the peak level of cases by 86.6%.

### Confirmed state-level caseload had only a small effect on social distancing timing

Controlling for partisanship, the number of confirmed COVID-19 cases in a state had only a small effect on the implementation of social distancing measures. (We emphasize this measure captures state-level cases: the total national cases are captured non-parametrically by the baseline hazard.) To show just how small this effect is, we compare a state with just four confirmed cases (the median among all state-days in our study period) to a state with 426 cases (the 95th percentile). The state with fewer cases was just 12.2% (95% CI: 6.5 to 17.8) less likely to implement social distancing on a given day, for a cumulative delay of just 0.38 days (95% CI: 0.36 to 0.41). While many of the early epicenters were Democratic-leaning states, it does not appear that the more aggressive action of Democratic states is a simple function of caseload.^5^ This is, from a public health perspective, a fairly reasonable result: early implementation of social distancing is likely to be far more effective, so pro-active governors may have realized there was little sense in waiting for a certain threshold of cases to act, especially given delays and failures in testing.

### Poorer states are less likely to adopt social distancing policies

All else equal, states at the 25th percentile of gross state product per capita were 26.2% (95% CI: 5.8 to 42.8) less likely to implement social distancing than states at the 75th percentile, which translates to an average delay of 0.90 days (95% CI: 0.83 to 0.95). This might explain Louisiana’s relatively slow adoption of social distancing policies despite a confirmed outbreak and a Democratic governor. Social distancing delays due to limited state economic resources are particularly troubling, as poorer states may also have larger vulnerable populations and more fragile health systems.

### Neighboring state actions increase the likelihood of social distancing policy

As in other policy areas, governors may look to their peers to determine appropriate action. The policy diffusion literature suggests different ways to determine each state’s “peer group.” Because the coronavirus spreads spatially, we focus on actions by neighboring states, and estimate the likelihood a state will implement a specific kind of social distancing as a function of the percentage of contiguous states also mandating that policy. We find that a state with no neighbors adopting a given policy was 32.0% (95% CI: 8.4 to 50.8) less likely to adopt it than a state with 50% or more of its neighbors adopting that policy. This corresponds to a delay of 1.20 days (95% CI: 1.13 to 1.26). Investigation of other possible mechanisms – including policy diffusion based on the number of confirmed COVID-19 cases in neighboring states or based on the actions of peers a given state typically looks to for policy innovations – failed to uncover additional patterns of diffusion. We speculate that actions by neighboring states may grant cover to governors to take similar actions.(28)

## Discussion

In this paper, we focused on decisions by governors, who through the structure of US and state constitutions and the abdication of presidential leadership found themselves on the frontlines of the battle against COVID-19. Future research should consider the role of local governments, but in March 2020, it was often governors who led their states – or allowed them to fall behind.

Why do Republican governors and states with Republican voters seem to resist social distancing policy, even controlling for many potential confounders? We strongly believe that realistic assessments of decision-making by elected officials must take electoral motivations and career ambitions seriously – as impolitic as that may be. Elected officials, regardless of party, must be responsive to the concerns of their voters and party leaders. However this essential feature of democratic representation does not inevitably produce the best policy outcome. Where the early stages of the COVID-19 pandemic were concerned, Republicans had fewer incentives to act quickly than Democrats. First, partisans of any stripe tend to minimize failures by their own party leaders and exaggerate failures by the opposing party.(34; 35; 36; 37) As a result, Republican voters took COVID-19 less seriously in the early stages of the US pandemic. Indeed, Republican voters were more concerned about Ebola during Obama’s presidency than they were about COVID-19 under Trump.(38) Second, all humans have difficulty accurately assessing risks that are small but not zero.(39; 40) In this case, Republican voters were more likely than Democrats to incorrectly assume that the COVID-19 fatality rate and disease burden was effectively the same as the flu.(15) Third, partisans’ worldviews are shaped by cues provided by their leaders.(34) In the early stages of the US pandemic, President Trump strongly signaled in press conference and on social media that the coronavirus was an exaggerated threat or even a hoax, a position that was magnified and reinforced by Republican-leaning media outlets. Finally, Republican officials almost surely considered the possibility of reprisals – both to their political careers and discretionary federal assistance to their states – from a president who frequently attacks members of his own party that he perceives to be disloyal.(41; 42; 43) To be clear, we are not arguing that politics was the sole motivation behind governors’ social distancing decisions: our results suggest otherwise. However, we do believe that the political headwinds were significantly greater for Republican Governors in Republican leaning states.

It is likely that governors will face this difficult decision again. Governors may choose to roll back social distancing at different points in time. Some experts contend that authorities will need to alternate between two months of social distancing and one month of restored freedoms to reduce the likelihood that health systems are overwhelmed by severe cases until a vaccine is developed, produced, and circulated.(3; 44) This implies state governments will be asked to impose difficult social distancing policies not once, but many times. Each instance will be a test of whether states can act promptly to prevent COVID-19 cases from peaking at unmanageable levels. If Republican governors and states with Republican majorities continue to lag behind, the cumulative impact on those states, and on the country as whole through spillovers, could be vast.

## Data Availability

Originally collected data on state-level social distancing policies can be obtained at http://covid19statepolicy.org/

## Appendix

**Table 1.**
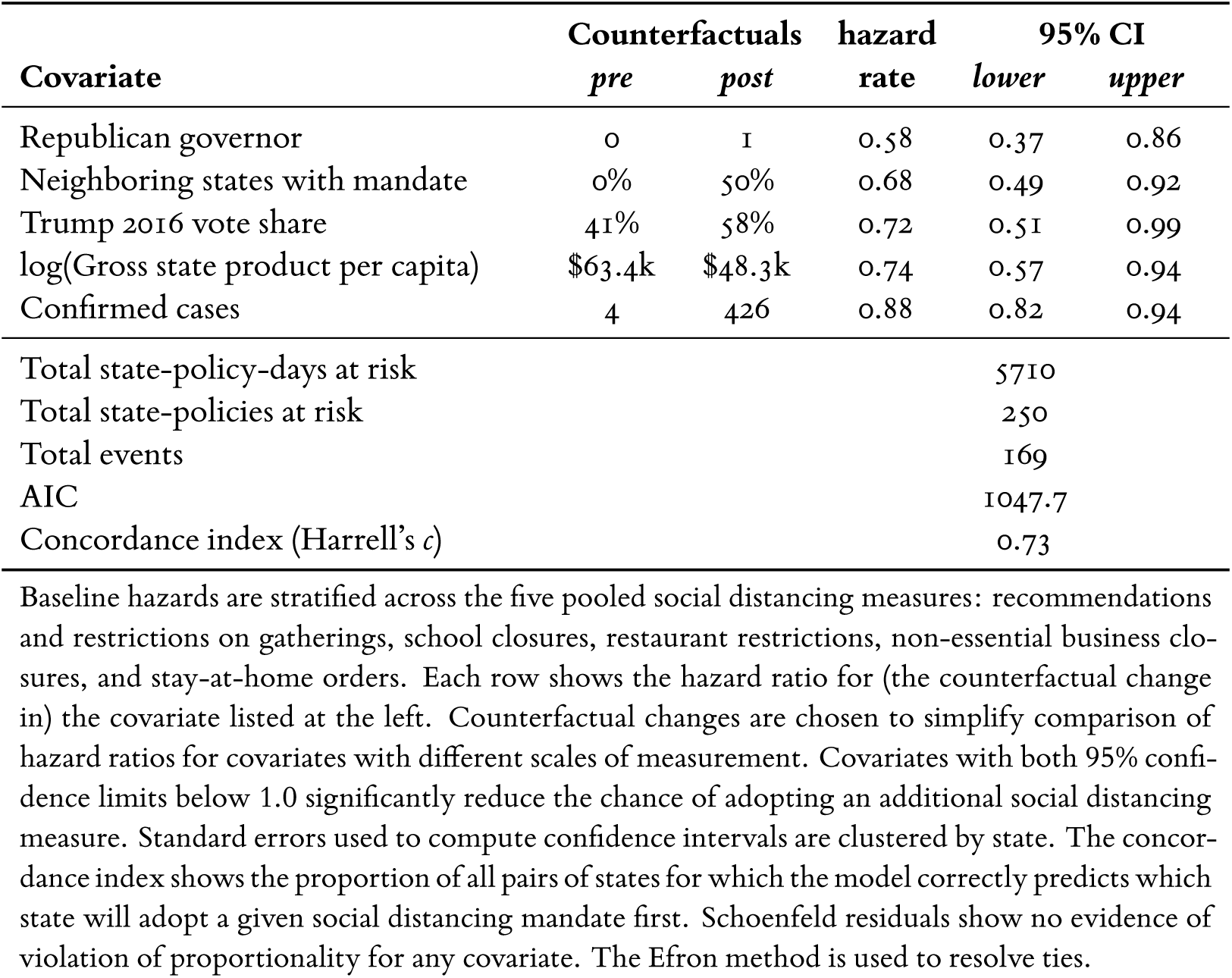
Pooled, stratified Cox proportional hazards model of state-level social distancing mandates, 26 February to 23 March 2020.

## Footnotes

1 We consider the first community transmission in the US more epidemiologically and politically relevant than initial reported transmission in each state. At the time of first community transmission in the US, virologists including Trevor Bedford suggested this strain had been circulating in Washington state since mid-January.(19) Given scarce and unreliable testing, governors faced a choice to either take immediate action as national cases increased or wait for (potentially belated) reports of in-state cases. We prefer to treat the decision to wait for a confirmed case as something to be explained, and so control for confirmed in-state cases in our model.

2 We include recommendations in this category for two reasons: first, this was often the first action states took, and early actors did not always revisit these policies with a restrictive man-date; second, the target of these measures was typically large events that would be unlikely to maintain viable attendance under the cloud of a state recommendation to cancel events.

3 R replication code sufficient to produce all figures can be found at [TO BE PROVIDED]. measures, including whether the state had ten or more confirmed cases, but found substantively similar results.

4 That is, we use the estimated Cox model to simulate the average of the marginal effects of each covariate on the timing of adopting each social distancing measure across the fifty states, and bootstrap confidence intervals around these marginal effects.(33)

5 The partisan result does not change if we consider simple transformations, such as whether the state had at least ten confirmed cases, or whether the state had a recorded death, or simply the total COVID-19 deaths; nor did any of these variables substantially explained social distancing adoption.

